# Integrating Clinical, Genetic, and Electrocardiogram-Based Artificial Intelligence to Estimate Risk of Incident Atrial Fibrillation

**DOI:** 10.1101/2024.08.13.24311944

**Authors:** Shinwan Kany, Joel T. Rämö, Samuel F. Friedman, Lu-Chen Weng, Carolina Roselli, Min Seo Kim, Akl C. Fahed, Steven A. Lubitz, Mahnaz Maddah, Patrick T. Ellinor, Shaan Khurshid

## Abstract

**Background:** AF risk estimation is feasible using clinical factors, inherited predisposition, and artificial intelligence (AI)-enabled electrocardiogram (ECG) analysis.

**Objective:** To test whether integrating these distinct risk signals improves AF risk estimation.

**Methods:** In the UK Biobank prospective cohort study, we estimated AF risk using three models derived from external populations: the well-validated Cohorts for Aging in Heart and Aging Research in Genomic Epidemiology AF (CHARGE-AF) clinical score, a 1,113,667-variant AF polygenic risk score (PRS), and a published AI-enabled ECG-based AF risk model (ECG-AI). We estimated discrimination of 5-year incident AF using time-dependent area under the receiver operating characteristic (AUROC) and average precision (AP).

**Results:** Among 49,293 individuals (mean age 65±8 years, 52% women), 825 (2.4%) developed AF within 5 years. Using single models, discrimination of 5-year incident AF was higher using ECG-AI (AUROC 0.705 [95%CI 0.686-0.724]; AP 0.085 [0.071-0.11]) and CHARGE-AF (AUROC 0.785 [0.769-0.801]; AP 0.053 [0.048-0.061]) versus the PRS (AUROC 0.618, [0.598-0.639]; AP 0.038 [0.028-0.045]). The inclusion of all components (“Predict-AF3”) was the best performing model (AUROC 0.817 [0.802-0.832]; AP 0.11 [0.091-0.15], p<0.01 vs CHARGE-AF+ECG-AI), followed by the two component model of CHARGE-AF+ECG-AI (AUROC 0.802 [0.786-0.818]; AP 0.098 [0.081-0.13]). Using Predict-AF3, individuals at high AF risk (i.e., 5-year predicted AF risk >2.5%) had a 5-year cumulative incidence of AF of 5.83% (5.33-6.32). At the same threshold, the 5-year cumulative incidence of AF was progressively higher according to the number of models predicting high risk (zero: 0.67% [0.51-0.84], one: 1.48% [1.28-1.69], two: 4.48% [3.99-4.98]; three: 11.06% [9.48-12.61]), and Predict-AF3 achieved favorable net reclassification improvement compared to both CHARGE-AF+ECG-AI (0.039 [0.015-0.066]) and CHARGE-AF+PRS (0.033 [0.0082-0.059]).

**Conclusions:** Integration of clinical, genetic, and AI-derived risk signals improves discrimination of 5-year AF risk over individual components. Models such as Predict-AF3 have substantial potential to improve prioritization of individuals for AF screening and preventive interventions.

## Introduction

Atrial fibrillation (AF) is associated with increased risks of stroke, heart failure, and death.^1^ AF screening can facilitate earlier diagnosis, and preventive treatment for AF-related morbidity, such as risk factor management or anticoagulation, can mitigate AF-related morbidity or even prevent AF altogether.^2^ However, screening approaches to date have demonstrated modest yield of AF diagnosis and have failed to demonstrate improvements in hard outcomes such as stroke or mortality.^3–5^

Analogous to established screening approaches for selected conditions such as lung cancer^6^ or osteoporosis,^7^ the efficiency of AF screening may be improved by utilizing a risk-informed approach. AF risk can be predicted with reasonable accuracy on the basis of clinical risk factors,^8^ inherited predisposition as assessed by polygenic risk scores [PRS]),^9,10^ and most recently artificial intelligence-enabled analysis of the electrocardiogram (ECG-AI).^11,12^ We have previously shown that the predictive power of a validated clinical risk score such as CHARGE-AF can be improved by the addition of either an AF PRS^9^ or ECG-AI.^11^ However, the degree to which each of these varied AF risk signals may overlap or complement one another within the context of a single comprehensive model remains unknown.

Here, we leveraged the UK Biobank – a unique resource of nearly 50,000 individuals with linkage to national health-related datasets, protocolized prospectively acquired 12-lead ECG, and genome-wide genotyping, to quantify the relative contributions of externally developed AF risk scores comprising a) clinical risk factors, b) common genetic variation, and c) AI-enabled ECG analysis. We hypothesized that by integrating complementary information, models incorporating each of the varying AF risk signals would achieve greater longitudinal AF discrimination.

## Methods

The UK Biobank was approved by the UK Biobank Research Ethics Committee (reference 11/NW/0382). All UK Biobank participants provided written informed consent. Use of UK Biobank (application #7089) data was approved by the MGB institutional review board.

### Data availability

UK Biobank data are accessible to researchers by application (www.ukbiobank.ac.uk). The PRS used in this study was obtained from the most recent AFGen consortium genome-wide association study by Roselli et al., which utilized a version of the analysis excluding UK Biobank participants.^13^ The published version of the ECG-AI model^11^ used to generate the AI-based predictions evaluated in the current analysis is available at https://github.com/broadinstitute/ml4h/tree/master/model_zoo/ECG2AF. Data processing scripts underlying the current analysis are available at https://github.com/shaankhurshid/af_prediction3.

### UK Biobank cohort

We analyzed the UK Biobank, a prospective cohort of 502,629 participants aged 40-69 that were recruited between 2006-2010.^14^ Participants underwent an extensive assessment in various stages (“instances”), including questionnaire data, anthropometric measures, and laboratory values. Instance 2 included a structured imaging visit for a subset of participants that included whole-body magnetic resonance imaging as well as protocolized resting 12-lead ECG. We analyzed individuals without prevalent AF who underwent resting 12-lead ECG at the instance 2 study visit.

### AF polygenic risk score

Details of genotyping, imputation, and quality control have been published previously and are provided in **Supplementary Methods**.^14,15^ The SNP-level weights used to calculate the AF PRS used in the current study were derived from a recent AF GWAS meta-analysis including 154,330 AF cases and 999,609 controls.^13^ The weights were derived using the PRS-CS approach.^16^ The UK Biobank cohort was not included in the version of the meta-analysis used as input to the PRS-CS algorithm. We calculated AF genetic risk in UK Biobank individuals using Plink2, summing up the weighted effect allele dosage of 1,113,667 variants included in the score with good quality (imputation info ≥0.4).^17^

### ECG-AI

To obtain ECG-based AI-enabled AF risk estimates, we applied a contemporary version of ECG-AI, a previously published convolutional neural network-based deep learning model designed to estimate 5-year risk of AF using a single 12-lead ECG.^11^ The current version of ECG-AI utilized a larger training set (450,000 standard 12-lead ECGs representing over 100,000 primary care and cardiology patients at Massachusetts General Hospital) and achieved higher discrimination of 5-year incident AF (c-index 0.761 vs. 0.716), but otherwise had identical architecture to the published model described in detail previously.^11^ UK Biobank participants were not included in any aspect of ECG-AI development.

### CHARGE-AF

To estimate the predictive utility of clinical risk factors, we calculated the CHARGE-AF score, a widely validated risk factor-based AF prediction tool.^8,18^ UK Biobank participants were not included in the original derivation of CHARGE-AF. To calculate the score, age, sex, race, height, weight, and blood pressure values were obtained from structured assessments using the value closest to the ECG date. Anti-hypertensive use was determined using self-reported medication data. Tobacco use was categorized as present or absent. Race was classified as White or non-White, as performed previously using CHARGE-AF. In cases of multiple available values, values from the assessment most closely preceding the ECG were used preferentially. The presence of heart failure, diabetes, and myocardial infarction were ascertained using previously published diagnostic and procedural codes.^9,11^ A full summary of clinical factor definitions is provided in **Supplementary Table 1**.

### Outcomes

The prediction target for each model was 5-year incident AF. AF events were identified using a previously published combination of self-reported illness codes, OPCS Classification of Interventions and Procedures version 4 codes for cardioversion or catheter ablation, and International Classification of Diseases, Ninth-or Tenth Revision (ICD-10) codes for AF or atrial flutter (**Supplementary Table 1**).^19^

### Statistical analysis

Discrimination of 5-year incident AF was measured using the area under the time-dependent receiver operating characteristic curve (AUROC) and time-dependent average precision (AP).^20,21^ Similar to how AUROC provides a composite measure of test sensitivity and specificity across a range of predictor thresholds, AP provides a composite measure of test precision (i.e., positive predictive value) and recall (i.e., sensitivity) across a range of predictor thresholds. Both AP and AUROC were calculated using inverse probability of censoring weights, which more accurately account for the potential bias introduced by censoring when compared to unweighted measures such as Harrell’s c-index.^22^

Discrimination of individual models was assessed as raw scores (CHARGE-AF, PRS) or probabilities (ECG-AI). Combination models were then developed by fitting Cox proportional hazards models with terms for each respective component (e.g., CHARGE-AF + PRS comprises a Cox model including a term for CHARGE-AF and a term for the AF PRS). As performed with the two-component Predict-AF model previously,^23^ prior to inclusion in Cox models the ECG-AI AF probabilities were logit-transformed to achieve an approximately linear relationship with log hazard. A model with a coefficient for each of CHARGE-AF, the PRS, and ECG-AI was termed “Predict-AF3”. Discrimination of the combination models was assessed using their linear predictors (i.e., score values). Discrimination was assessed overall and across subgroups of age (age<60 years, age 60-70 years, age>70 years, approximating tertiles of the distribution) and sex.

We then assessed model calibration. To allow fair comparison across each component, including components with no intrinsic translation to a longitudinal risk estimate (i.e., AF PRS), we fit univariable Cox proportional hazards models with a single term for each individual component. Linear predictors of the individual fitted models as well as the combination models outlined above were then converted into 5-year absolute risk estimates using the equation 1−S_0_^exp(ΣX−ΣY)^, where S_0_ is the average 5-year AF-free survival of the sample, ΣX is the individual’s linear predictor or score value, and ΣY is the average score of the sample. Calibration of absolute risk estimates was then assessed by plotting smoothed curves of absolute versus predicted risk using adaptive hazard regression, and calculating the integrated calibration index (ICI), a measure of the average prediction error weighted by the empirical risk distribution and where an ICI of zero indicates perfect absolute risk estimates.^24^ For AP, AUROC, and ICI estimates, 95% confidence intervals were calculated using bootstrapping (500 iterations for AP and AUROC, and 200 iterations for ICI), which also provided estimates of standard error to perform pairwise *z* testing. To estimate the clinical impact of implementing one AF risk model over another, we calculated net reclassification improvement, both at the >2.5% 5-year AF risk threshold and as a continuous measure.^8,25^ We considered 2-sided p<0.05 statistically significant. Analyses were performed using Python v3.10 and R v4.3.

## Results

### Sample *characteristics*

We analyzed 49,293 individuals without prevalent AF (age 65 ± 8 years, 52% women, 96% White) at the time of the UK Biobank instance 2 visit. Over 5 years of follow-up, 825 participants (2.4%) developed incident AF. An overview of the study is provided in **Figure 1** and baseline characteristics are provided in **Table 1**.

**Figure 1.**
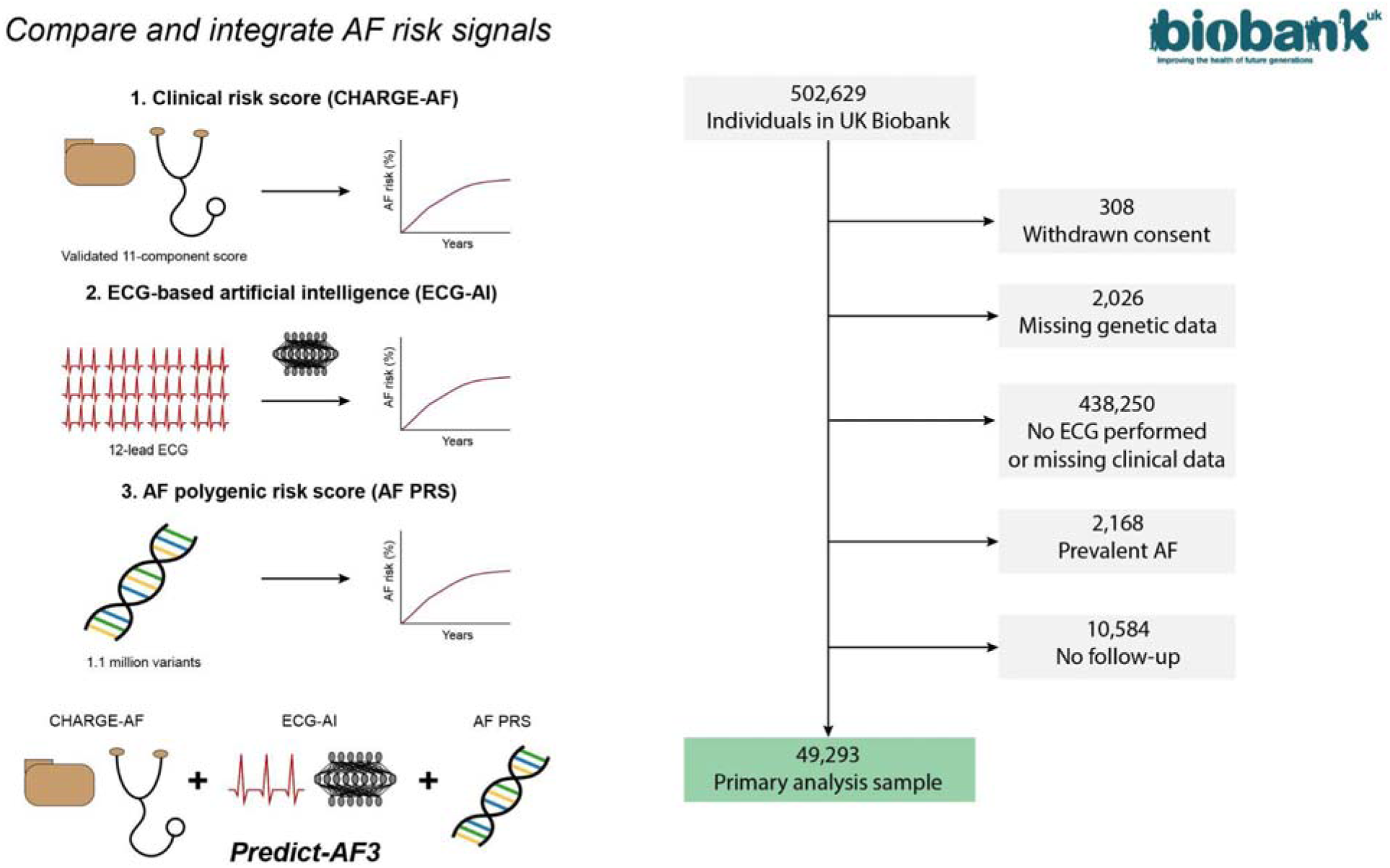
Flow chart of study cohort Depicted is an overview of the cohort creation. CHARGE-AF is a clinical risk model for incident atrial fibrillation (AF) risk prediction. ECG-AI is a deep-learning model trained to predict 5-year risk of AF and the polygenic risk score (PRS) is a genetic risk score estimating the risk based on common variant risk from genome-wide association studies of incident AF. Predict-AF3 combines all three risk prediction models for a comprehensive AF risk prediction.

**Table 1.** Baseline Characteristics.

|  | Female | Male | Overall |
| --- | --- | --- | --- |
| N | 25742 | 23551 | 49293 |
| Age at ECG, years | 64.48 (7.7) | 65.73 (7.9) | 65.08 (7.8) |
| Height, cm | 162.6 (6.3) | 175.9 (6.6) | 169.0 (9.2) |
| Weight, kg | 69.1 (13.4) | 83.7 (13.6) | 76.1 (15.3) |
| Current smoker | 762 (3.0) | 971 (4.1) | 1733 (3.5) |
| Systolic blood pressure, mmHg | 136.70 (19.7) | 142.53 (17.6) | 139.49 (18.9) |
| Diastolic blood pressure, mmHg | 79.25 (10.1) | 77.66 (10.0) | 80.99 (9.9) |
| Diabetes mellitus | 1136 (4.4) | 1928 (8.2) | 3064 (6.2) |
| Heart failure | 80 (0.3) | 173 (0.7) | 253 (0.5) |
| Myocardial infarction | 235 (0.9) | 952 (4.0) | 1187 (2.4) |
| Values are shown as mean (standard deviation) for continuous measures and N (%) for categorical measures. |  |  |  |

### Discrimination of incident atrial fibrillation using single models

Using single prediction models, discrimination of 5-year incident AF as measured by AUROC was highest using CHARGE-AF (0.785 [95% CI: 0.769-0.801]), followed by ECG-AI 0.705 [0.686-0.724]) and the PRS (0.618 [0.598-0.639]). Discrimination using CHARGE-AF tended to increase with longer prediction windows, while discrimination using ECG-AI and AF PRS remained largely stable over time (**Figure 2**). While CHARGE-AF had highest discrimination among individuals aged <60 years (AUROC 0.739 [0.678-0.798]), discrimination was more similar across models in those aged >70 years (e.g. CHARGE-AF AUROC 0.688 vs ECG-AI AUROC 0.687) (**Supplementary Table 2**). Models performed similarly among men versus women (**Supplementary Table 3**).

**Figure 2.**
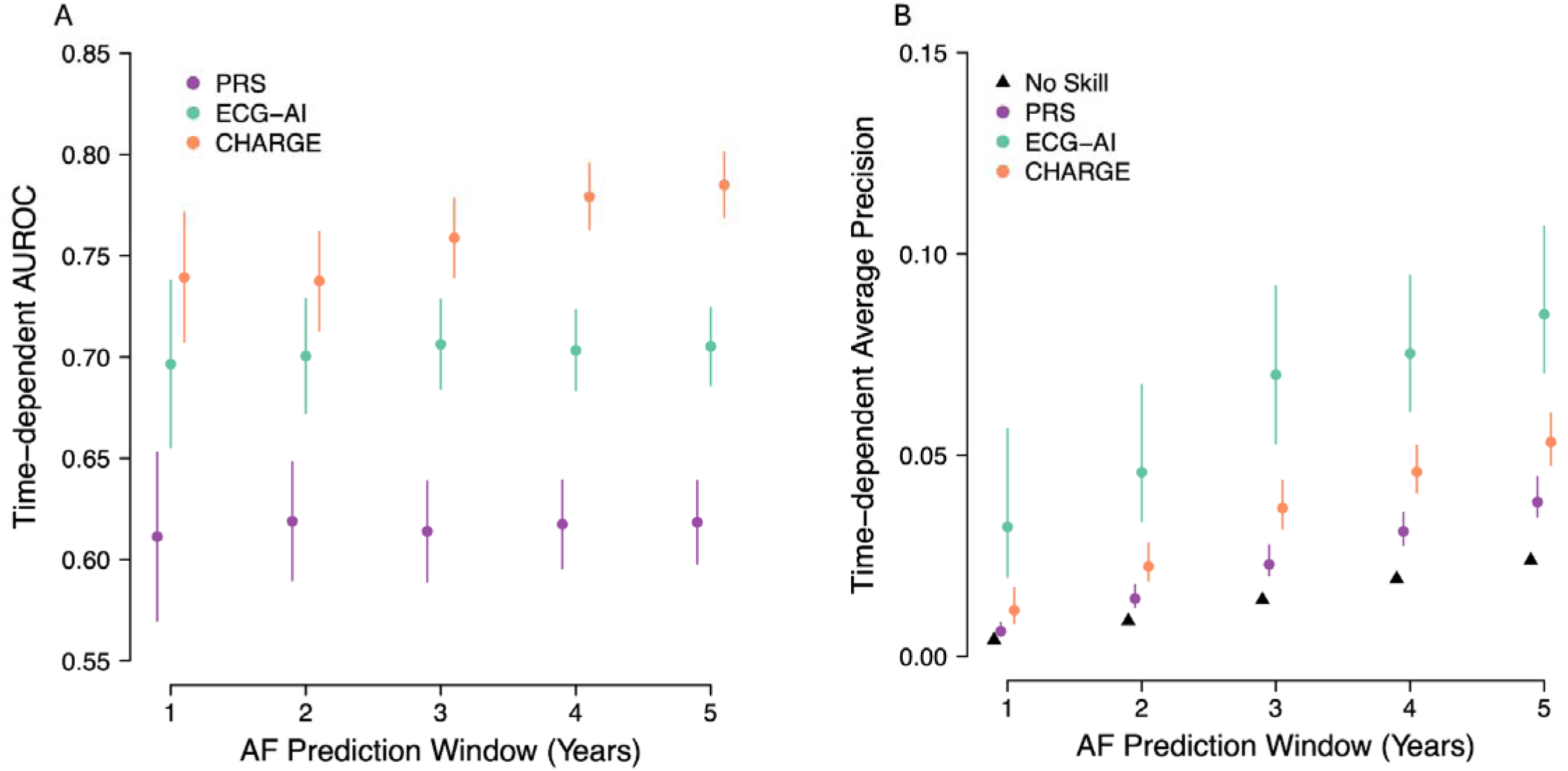
Discrimination of incident atrial fibrillation Depicted is model discrimination for a 5-year window of incident atrial fibrillation (AF) using a polygenic risk score (PRS) model (teal), an ECG-derived artificial intelligence (AI) prediction (turquoise) and a model based on the CHARGE-AF score (orange). Panel A depicts discrimination measured using a time-dependent area under the receiver operating characteristic curve (AUROC) while Panel B depicts the time-dependent average precision. A model with no discriminative power is depicted by black triangles.

Measured using average precision, discrimination was highest using ECG-AI (0.085 [0.071-0.11]), followed by CHARGE-AF (0.053 [0.048-0.061]) and the PRS (0.038 [0.028-0.045]), indicative of relatively good performance using ECG-AI for the detection of the highest risk individuals. Consistent with an increasing cumulative event rate (e.g., 0.4% at 1 year to 2.4% at 5 years), the AP for each model increased over time (**Figure 2**) and among older subgroups of age (**Supplementary Table 2**). Trends in performance were similar among men versus women (**Supplementary Table 3**).

As expected, there was no substantive correlation between PRS and CHARGE-AF (r=-0.014 [-0.022 to -0.0048]). There was very weak correlation between PRS and ECG-AI (r=0.041 [0.032-0.050]). There was moderate correlation between ECG-AI and CHARGE-AF (r=0.34 [0.33-0.35])

### Discrimination of incident atrial fibrillation using combined models

We then evaluated discrimination of the combined models, namely a) CHARGE-AF + PRS, b) ECG-AI + PRS, c) CHARGE-AF + ECG-AI, and CHARGE-AF + ECG-AI + PRS (“Predict-AF3”). The best performing two-component models were CHARGE-AF + ECG-AI (AUROC 0.802 [0.786-0.818]; AP 0.098 [0.081-0.13]), followed by CHARGE-AF + PRS (AUROC 0.802 [0.786-0.818]; AP 0.053 [0.048-0.061]). The best performing model overall was Predict-AF3 (AUROC 0.817 [0.802-0.832]; AP 0.11 [0.091-0.15], p<0.01 vs CHARGE-AF + ECG-AI and CHARGE-AF + PRS) (**Figure 3**). Addition of the PRS to each component contributed relatively modest but consistently detectable improvements in discrimination (improvement in AUROC with addition of PRS to CHARGE-AF: 0.017 [0.0096-0.025], addition of PRS to ECG-AI: 0.023 [0.012-0.034], addition of PRS to CHARGE-AF + ECG-AI: 0.015 [0.0089-0.021]) (**Supplementary Table 4**). Improvements were similar according to AP (**Supplementary Table 4**). Relative performance patterns were similar when assessing discrimination of AF at 1 year (e.g., Predict-AF3 AUROC 0.791 [0.758-0.823]; AP 0.030 [0.020-0.050], **Figure 3**).

**Figure 3.**
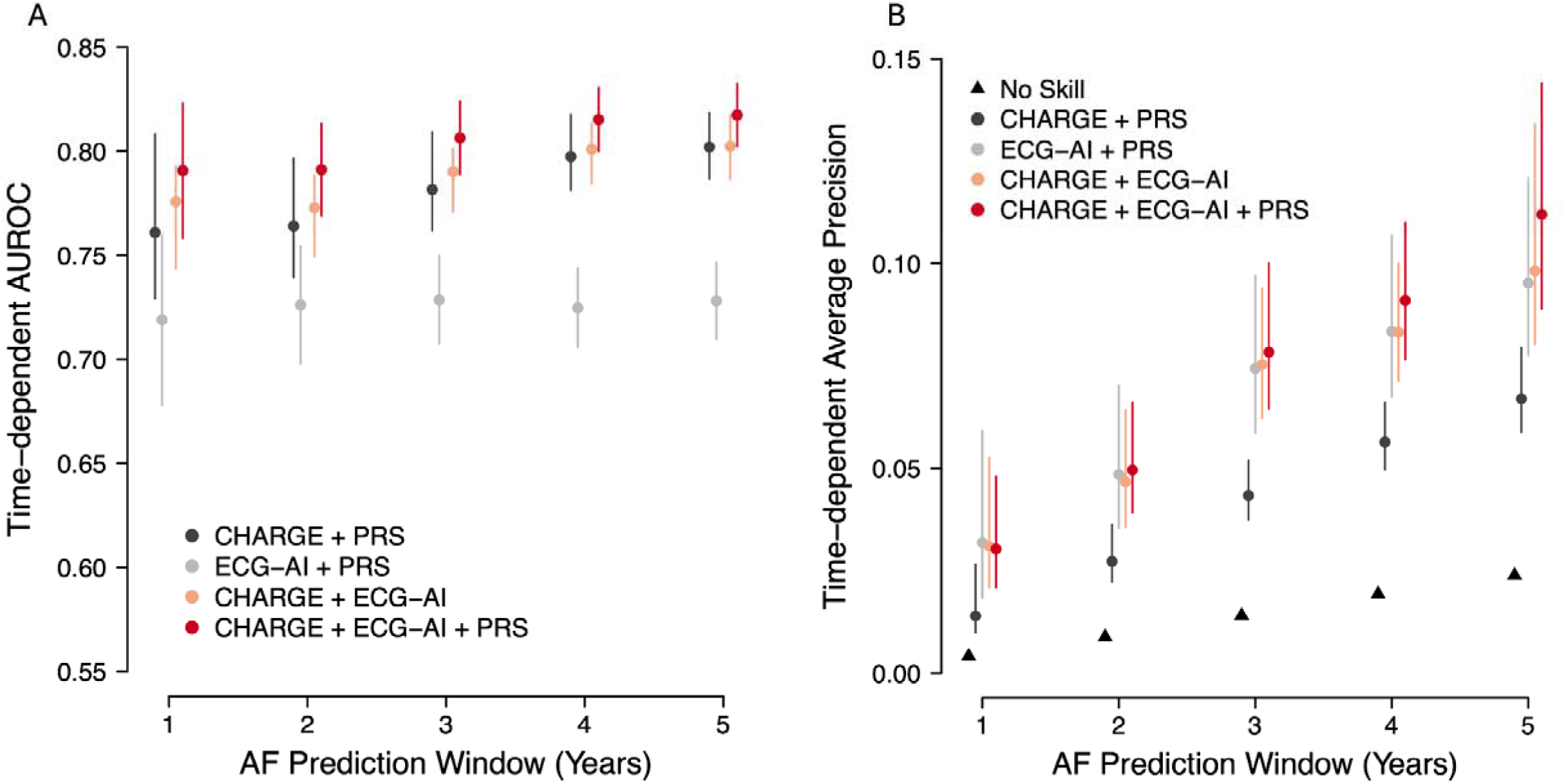
Discrimination of incident atrial fibrillation in combined models Depicted is model discrimination for a 5-year window of incident atrial fibrillation (AF) using models a) combining polygenic risk score (PRS) and CHARGE-AF (black), a model combining an ECG-derived artificial intelligence (ECG-AI) prediction and the PRS (grey), a model combining ECG-AI and the CHARGE-AF (orange), and a model combining ECG-AI, CHARGE-AF and the PRS (red). Panel A depicts discrimination measured using a time-dependent area under the receiver operating characteristic curve (AUROC) while Panel B depicts the time-dependent average precision. A model with no discriminative power is depicted by black triangles.

### Calibration and net reclassification of atrial fibrillation risk estimates

Absolute risk estimates for the single and combined models were well-calibrated (**Supplementary Figure 1**). Absolute error rates were consistently very low (ICI range 4.16x10^-5^ for CHARGE-AF to 0.0018 for Predict-AF3). At the >2.5% 5-year AF risk threshold, Predict-AF3 resulted in favorable net reclassification improvement versus CHARGE-AF + ECG-AI (NRI 0.039 [0.015-0.066]), which was driven by both favorable case reclassification (NRI+ 2.59% [0.18-5.30]) and favorable non-case reclassification (NRI-1.35% [1.06-1.67]) (**Supplementary Table 5**). Predict-AF3 also provided similar reclassification improvement over CHARGE-AF + PRS, although it was driven solely by favorable non-case reclassification (NRI 0.033 [0.0082-0.059]; NRI+ 0.52% [-2.06 to 3.00]; NRI-2.81% [2.54-3.08]) (**Supplementary Table 5**). Continuous reclassification improvement was also favorable using Predict-AF3 (0.34 [0.26-0.41] vs. CHARGE-AF + ECG-AI; 0.23 [0.16-0.31] vs. CHARGE-AF + PRS).

### Stratification of longitudinal atrial fibrillation incidence using Predict-AF3

Use of Predict-AF3 effectively stratified longitudinal AF risk, with individuals at high AF risk (5-year predicted AF risk >2.5%) having a 5-year cumulative incidence of AF of 5.83% (5.33-6.32), those with a predicted risk of 1-2.5% having a cumulative incidence of 1.51% (1.26-1.75) and those with a predicted risk of ≤1% having a cumulative incidence of 0.56% (0.43-0.69) (**Figure 4**).

**Figure 4.**
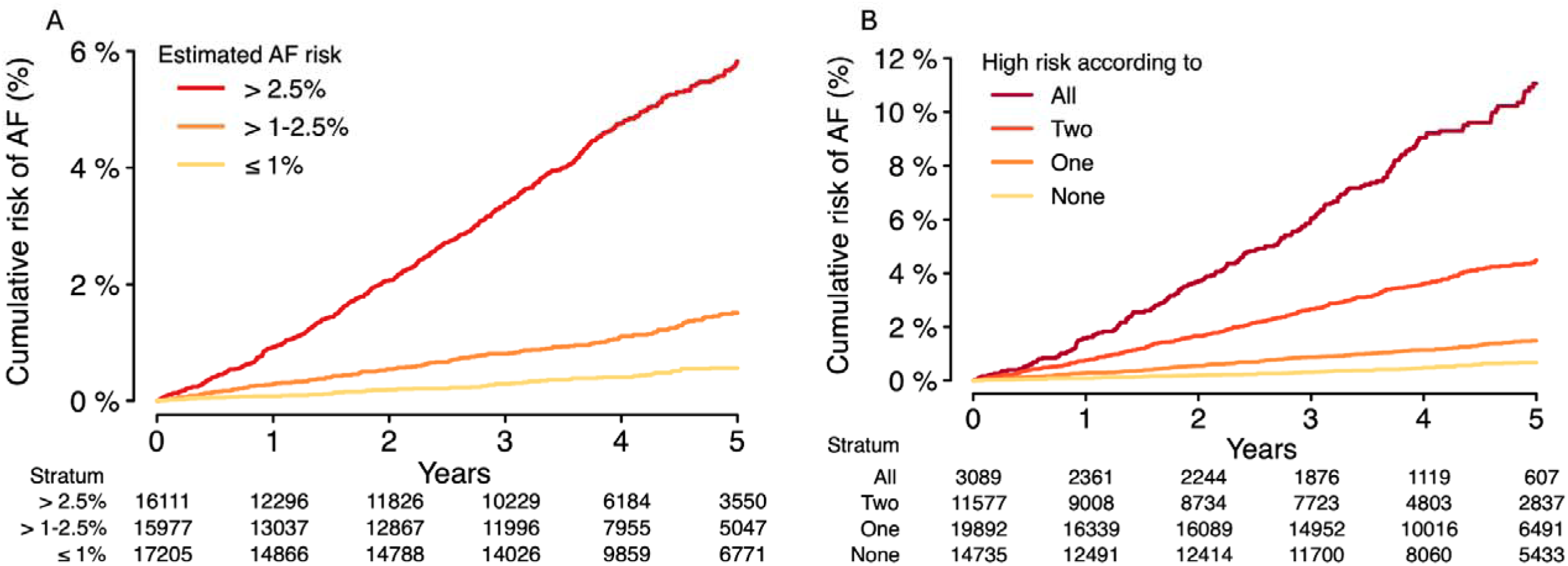
Cumulative risk of incident atrial fibrillation stratified by the combined models Depicted is the cumulative risk of AF across strata of predicted risk using Predict-AF3. Panel A plots cumulative risk across categories of Predict-AF3 estimated risk (thresholds chosen to approximate tertiles of the risk distribution), and Panel B plots cumulative risk across strata of high risk (i.e., 2-year AF risk >2.5%) by each model component. The number at risk across each stratum over time is depicted below each plot.

Using the >2.5% 5-year AF risk threshold, the 5-year cumulative incidence of AF was progressively higher according to the number of individual models predicting high risk (zero: 0.67% [0.51-0.84], one: 1.48% [1.28-1.69], two 4.48% [3.99-4.98], three: 11.06% [9.48-12.61]) (**Figure 4**). Among the 825 participants who developed AF during the 5-year period, 208 (25.2%) had high estimated AF risk according to all three models, 340 (41.2%) according to two models, 207 (25.1%) according to one model, and 70 (8.5%) according to zero models.

## Discussion

In this study, we leveraged a unique resource of nearly 50,000 prospective cohort study participants with detailed clinical data, genome-wide genotyping, and protocolized 12-lead ECG to compare the relative predictive utility of varied forms of AF risk information. Specifically, we applied externally derived and contemporary clinical, genetic and ECG-based AI-enabled AF risk models separately and in combination. Our findings demonstrate that clinical, genetic, and AI-based AF risk signals are complementary. There was a graded increase in AF incidence as individuals were identified as high-risk by a greater number of risk signals. The Predict-AF3 score, which combines all three elements, achieved higher predictive utility than any single or two-model combination. Overall, our work establishes the value of integrating varying data types to achieve increasingly accurate AF risk estimates, providing a foundation for efforts to better prioritize individuals for AF screening and related preventive efforts. Our findings yield several key implications.

First, it is feasible to integrate varying AF risk signals to achieve more accurate AF risk estimates. In our analysis, the Predict-AF3 score consistently exhibited favorable discrimination compared to the individual components of the score, implying that clinical, genetic, and ECG-based AI risk estimates provide complementary information. By incorporating all three components, Predict-AF3 achieves the highest discrimination of 5-year incident AF risk (AUROC 0.82, AP 0.11) of any AF prediction model previously applied in a prospective community cohort.^8,9,11,18,26,27^ Our findings are consistent with prior observations from our group suggesting that the addition of ECG-derived AI risk to clinical risk improves AF risk estimation.^11^ Despite some evidence that ECG-based AI may encode some aspects of inherited predisposition to AF,^28^ we consistently observed improvements in AF discrimination with the addition of PRS to any individual component or two-component combination, suggesting that genetic risk remains orthogonal to current ECG-based AI models. However, as with prior work, the degree of discrimination improvement observed with the addition of PRS was generally modest,^9,29^ although we did note favorable net reclassification improvement when adding the PRS to the best performing two-component model (CHARGE-AF + ECG-AI) to create Predict-AF3. Importantly, we observed that AF incidence increased progressively with the number of distinct elements portending high AF risk, suggesting that individuals with risk conferred by the combination of clinical factors, genetics, and AI-enabled ECG signals appear particularly vulnerable to developing AF.

Second, the optimal approach to AF risk estimation to guide AF screening and related preventive interventions may be a function of available data and the specific setting in which risk stratification is intended. Although Predict-AF3 achieved the highest discrimination of longitudinal AF among all approaches tested, genome-wide genotyping data is not commonly available in many populations in which AF screening is considered (e.g., routine primary care). To this end, we observed good discrimination using CHARGE-AF + ECG-AI, a model requiring only routine clinical factors and a single 12-lead ECG, an inexpensive diagnostic test available within minutes in most clinic settings. Future work is warranted to assess whether the predictive utility of ECG-AI may extend to single-lead ECGs, which are increasingly available using mobile and consumer devices and may increase the reach of AF risk estimation further.^30^ Conversely, the combination of CHARGE-AF + PRS also achieved similar performance to CHARGE-AF + ECG-AI. Such a model may have particular value in population health interventions targeting healthcare-related biobanks, where risk estimation can be run on previously acquired samples, and leveraging linked electronic health record data.

Third, the integration of an increasing variety of clinically relevant risk markers has potential to further the goal of personalized medicine with regard to AF. While opportunistic screening is recommended in the most recent European Society of Cardiology guidelines for people of older age,^31^ recent randomized trials of screening guided only by age have resulted in little to no increase in AF diagnosis, and absence of meaningful improvements in AF-related complications such as stroke or mortality.^3,4^ Subgroup analyses have suggested higher AF screening yield among individuals with higher AF incidence,^3^ suggesting that screening based on AF risk may be more efficient. To this end, recent work has identified the ability to stratify AF risk based on a large breadth of potential markers, including not only clinical factors, polygenic risk, and AI-based signals, but additionally imaging-based features and blood-based biomarkers.^32,33^ Therefore, although Predict-AF3 provides an important demonstration of the potential value of incorporating varying contributors to AF risk to achieve more precise AF risk estimates, future work is warranted to integrate even more markers, and potentially leverage emerging statistical learning techniques capable of incorporating high-level interactions between varied data types. Ultimately, randomized trials are needed to assess whether AF risk estimation using progressively more accurate models leads to more efficient AF screening interventions.

## Limitations

Our study should be interpreted in the context of design. The UK Biobank is primarily of European ancestry and our results might not be generalizable to people of other ancestries. Second, the UK Biobank is not reflective of the general population and comprises healthier individuals.^34^ Due to survivorship bias, the instance 2 cohort that received protocolized 12-lead ECG is even healthier than the UK Biobank at large. Third, our models were assessed against the clinical diagnosis of AF, and therefore their performance for detection of undiagnosed AF in a screening setting may vary. Fourth, although the clinical risk score, PRS, and ECG-based AI model assessed in the current study were all developed externally to the UK Biobank, the relative contributions of each component in the combination models were weighted within-sample. The relative importance of AF risk components may differ in other populations, and therefore further validation of the Predict-AF3 multi-component model is warranted.

## Conclusions

Integration of clinical, genetic, and ECG-based AI risk signals for AF into a single model (Predict-AF3) leads to greater predictive utility of 5-year incident AF compared to the use of individual elements. Consistent with the presence of complementary information, AF incidence increased progressively with the total number of distinct elements portending high AF risk. Scores such as Predict-AF3 may pave the way for integrated and personalized prioritization of individuals for AF screening and related preventive interventions.

## Data Availability

UK Biobank data are accessible to researchers by application (www.ukbiobank.ac.uk). The PRS used in this study was obtained from the most recent AFGen consortium genome-wide association study by Roselli et al., which utilized a version of the analysis excluding UK Biobank participants.13 The published version of the ECG-AI model11 used to generate the AI-based predictions evaluated in the current analysis is available at https://github.com/broadinstitute/ml4h/tree/master/model_zoo/ECG2AF. Data processing scripts underlying the current analysis are available at https://github.com/shaankhurshid/af_prediction3.

## Acknowledgments

This research has been conducted using the UK Biobank Resource under Application Number 17488. The authors would like to thank study staff and investigators from the UK Biobank.

## Authors’ Contributions

SKany – Study conception, design, data analysis, data interpretation, drafting

SKhurshid – Study conception, design, data analysis, data interpretation, drafting, critical review and editing

CR, LCW, SFF – Data analysis, data interpretation, critical review

PTE – Study conception, design, data interpretation, drafting, critical review

JTR, MSK, ACF, SAL AAP, MM – Data interpretation, critical review

