## Supplement for "Integrating Clinical, Genetic, and Electrocardiogram-Based Artificial Intelligence to Estimate Risk of Incident Atrial Fibrillation"

**Table of Contents**

**Supplementary methods	3**

Genotyping, imputation and quality control	3

**Supplementary Tables	4**

Table 1 Clinical factor definitions	4

Table 2 Predictive performance of models for 5-year atrial fibrillation risk by subgroups of age	13

Table 3 Predictive performance of models for 5-year atrial fibrillation risk by subgroups of sex	14

Table 4 Increase in discrimination for 5-year risk of incident atrial fibrillation	15

Table 5 Net reclassification of incident AF risk using Predict-AF3 versus two-component models	16

**Supplementary Figures	17**

Figure 1: Calibration of incident atrial fibrillation	17

**References	19**

### Supplementary methods

#### Genotyping, imputation and quality control

Nearly 488,000 participants in the UK Biobank were genotyped with either the UK BiLEVE or UK Biobank Axiom array and imputation conducted with the UK10K+1000 Genomes phase III and Haplotype Reference Consortium panel.(Bycroft et al. 2018) Both arrays were used to capture short insertions and deletions as well as single nucleotide polymorphism (SNPs) with a shared marker content of 95%. Quality control for these genotyped variants included removal of variants with minor allele frequency (MAF) less than 1%, genotyping call rate less than 90%, or Hardy-Weinberg violation within the GWAS participants at P <= 1E-15.

Sample-level and quality-control (QC) included removal of those without imputed data, sample-level genotype missingness greater than or equal to 2%, aneuploidy of sex chromosomes, outliers for heterozygosity, and excessive third-degree relatives as centrally computed by the UK Biobank.(Bycroft et al. 2018)

### Supplementary Tables

#### **Table 1 Clinical factor definitions**

| **Phenotype** | **Data fields** | **Field names** | **Data codes** | **Data code definitions** |
| --- | --- | --- | --- | --- |
| Atrial fibrillation | 20002 | Non-cancer illness code, self-reported | 1471, 1483 | Atrial fibrillation, Atrial flutter |
| Atrial fibrillation | 20004 | Operation code, self-reported | 1524 | Cardioversion |
| Atrial fibrillation | 41202  41204 40001 40002 | Diagnoses - main ICD10  Diagnoses – secondary ICD10  Underlying (primary) cause of death: ICD10  Contributory (secondary) cause of death: ICD10 | I48, I48.0, I48.1, I48.2, I48.3, I48.4, I48.9 | Atrial fibrillation and flutter, Paroxysmal atrial fibrillation, Persistent atrial fibrillation, Chronic atrial fibrillation, Typical atrial flutter, Atypical atrial flutter, Atrial fibrillation and flutter, unspecified |
| Atrial fibrillation | 41203  41205 | Diagnosis - main ICD9  Diagnoses - secondary ICD9 | 4273 | Atrial fibrillation and flutter |
| Atrial fibrillation | 41200  41210 | Operative procedures – main OPCS  Operative procedures – secondary OPCS | K57.1, K62.1, K62.2, K62.3, K62.4, X50.1, X50.2 | Percutaneous transluminal ablation of atrioventricular node, Percutaneous transluminal ablation of pulmonary vein to left atrium conducting system, Percutaneous transluminal ablation of atrial wall for atrial flutter, Percutaneous transluminal ablation of conducting system of heart for atrial flutter NEC, Percutaneous transluminal internal cardioversion NEC, Direct current cardioversion, External cardioversion NEC |
| Diabetes | 2443 | Diabetes diagnosed by doctor | 1 | Yes |
| Diabetes | 20002 | Non-cancer illness code, self-reported | 1220, 1221, 1222, 1223 | Diabetes, Gestational diabetes, Type 1 diabetes, Type 2 diabetes |
| Diabetes | 2986 | Insulin use within one year | 1 | Started insulin within one year diagnosis of diabetes - Yes |
| Diabetes | 6177 | Medication for cholesterol, blood pressure or diabetes | 3 | Insulin |
| Diabetes | 6153 | Medication for cholesterol, blood pressure, diabetes, or take exogenous hormones | 3 | Insulin |
| Diabetes | 41202  41204 40001 40002 | Diagnoses - main ICD10  Diagnoses – secondary ICD10  Underlying (primary) cause of death: ICD10  Contributory (secondary) cause of death: ICD10 | E10, E10.0, E10.1, E10.2, E10.3, E10.4, E10.5, E10.6, E10.7, E10.8, E10.9, E11, E11.0, E11.1, E11.2, E11.3, E11.4, E11.5, E11.6, E11.7, E11.8, E11.9, E12, E12.1, E12.8, E12.9, E13, E13.1, E13.2, E13.3, E13.5, E13.6, E13.7, E13.8, E13.9, E14, E14.0, E14.1, E14.2, E14.3, E14.4, E14.5, E14.6, E14.7, E14.8, E14.9 | Insulin-dependent diabetes mellitus, Insulin-dependent diabetes mellitus with coma, Insulin-dependent diabetes mellitus with ketoacidosis, Insulin-dependent diabetes mellitus with renal complications, Insulin-dependent diabetes mellitus with ophthalmic complications, Insulin-dependent diabetes mellitus with neurological complications, Insulin-dependent diabetes mellitus with peripheral circulatory complications, Insulin-dependent diabetes mellitus with other specified complications, Insulin-dependent diabetes mellitus with multiple complications, Insulin-dependent diabetes mellitus with unspecified complications, Insulin-dependent diabetes mellitus without complications, Non-insulin-dependent diabetes mellitus, Non-insulin-dependent diabetes mellitus - with coma, Non-insulin-dependent diabetes mellitus - with ketoacidosis, Non-insulin-dependent diabetes mellitus - with renal complications, Non-insulin-dependent diabetes mellitus - with ophthalmic complications, Non-insulin-dependent diabetes mellitus - with neurological complications, Non-insulin-dependent diabetes mellitus - with peripheral circulatory complications, Non-insulin-dependent diabetes mellitus - with other specified complications, Non-insulin-dependent diabetes mellitus - with multiple complications, Non-insulin-dependent diabetes mellitus - with unspecified complications, Non-insulin-dependent diabetes mellitus - without complications, Malnutrition-related diabetes mellitus, Malnutrition-related diabetes mellitus with ketoacidosis, Malnutrition-related diabetes mellitus with unspecified complications, Malnutrition-related diabetes mellitus without complications, Other specified diabetes mellitus, Other specified diabetes mellitus with ketoacidosis, Other specified diabetes mellitus with renal complications, Other specified diabetes mellitus with ophthalmic complications, Other specified diabetes mellitus with peripheral circulatory complications, Other specified diabetes mellitus with other specified complications, Other specified diabetes mellitus with multiple complications, Other specified diabetes mellitus with unspecified complications, Other specified diabetes mellitus without complications, Unspecified diabetes mellitus, Unspecified diabetes mellitus with coma, Unspecified diabetes mellitus with ketoacidosis, Unspecified diabetes mellitus with renal complications, Unspecified diabetes mellitus with ophthalmic complications, Unspecified diabetes mellitus with neurological complications, Unspecified diabetes mellitus with peripheral circulatory complications, Unspecified diabetes mellitus with other specified complications, Unspecified diabetes mellitus with multiple complications, Unspecified diabetes mellitus with unspecified complications, Unspecified diabetes mellitus without complications |
| Diabetes | 41203  41205 | Diagnosis - main ICD9  Diagnoses - secondary ICD9 | 2500, 25000, 25001, 25009, 2501, 25011, 25019, 2503, 2504, 2505, 25099 | Diabetes mellitus without mention of complication, Diabetes mellitus without mention of complication (adult-onset type), Diabetes mellitus without mention of complication (juvenile type), Diabetes mellitus without mention of compl. (adult/juvenile unspec.), Diabetes with ketoacidosis, Diabetes with ketoacidosis (juvenile type), Diabetes with ketoacidosis (adult/juvenile unspec.), Diabetes with renal manifestations, Diabetes with ophthalmic manifestations, Diabetes with neurological manifestations, Diabetes with unspecified complications (unspecified onset) |
| Heart Failure | 20002 | Non-cancer illness code, self-reported | 1076, 1079 | Heart failure/pulmonary oedema, Cardiomyopathy |
| Heart Failure | 41202  41204  40001  40002 | Diagnoses - main ICD10  Diagnoses – secondary ICD10  Underlying (primary) cause of death: ICD10  Contributory (secondary) cause of death: ICD10 | I11.0, I13.0, I13.2, I25.5, I42.0, I42.5, I42.8, I42.9, I50, I50.0, I50.1, I50.9 | Hypertensive heart disease with (congestive) heart failure, Hypertensive heart and renal disease with (congestive) heart failure, Hypertensive heart and renal disease with both (congestive) heart failure and renal failure, Hypertensive heart disease with (congestive) heart failure, Hypertensive heart and renal disease with (congestive) heart failure, Hypertensive heart and renal disease with both (congestive) heart failure and renal failure, Ischaemic cardiomyopathy, Dilated cardiomyopathy, Other restrictive cardiomyopathy, Other cardiomyopathies, Cardiomyopathy, unspecified, Heart failure, Congestive heart failure, Left ventricular failure, Heart failure, unspecified |
| Hypertension | 20002 | Non-cancer illness code, self-reported | 1065, 1072 | essential hypertension, hypertension |
| Hypertension | 41203  41205 | Diagnosis - main ICD9  Diagnoses - secondary ICD9 | 4019, 4020, 4021, 4029, 4030, 4031, 4039, 4040, 4041, 4049, 4050, 4051, 4059 | Essential hypertension, not specified as malignant or benign, Hypertensive heart disease, specified as malignant, Hypertensive heart disease, specified as benign, Hypertensive heart disease, not specified as malignant or benign, Hypertensive renal disease, specified as malignant, Hypertensive renal disease, specified as benign, Hypertensive renal disease, not specified as malignant or benign, Hypertensive heart and renal disease, specified as malignant, Hypertensive heart and renal disease, specified as benign, Hypertensive heart and renal disease, not specified as malignant or benign, Secondary hypertension, specified as malignant, Secondary hypertension, specified as benign, Secondary hypertension, not specified as malignant or benign |
| Hypertension | 41202  41204  40001  40002 | Diagnoses - main ICD10  Diagnoses – secondary ICD10  Underlying (primary) cause of death: ICD10  Contributory (secondary) cause of death: ICD10 | I10,I11,I11.0,I11.9,I12,I12.0,I12.9,I13,I13.0,I13.1,I13.2,I13.9,I15,I15.0,I15.1,I15.2,I15.8,I15.9 | Essential (primary) hypertension, Hypertensive heart disease, Hypertensive heart disease with (congestive) heart failure, Hypertensive heart disease without (congestive) heart failure, Hypertensive renal disease, Hypertensive renal disease with renal failure, Hypertensive renal disease without renal failure, Hypertensive heart and renal disease, Hypertensive heart and renal disease with (congestive) heart failure, Hypertensive heart and renal disease with renal failure, Hypertensive heart and renal disease with both (congestive) heart failure and renal failure, Hypertensive heart and renal disease, unspecified, Secondary hypertension, Renovascular hypertension, Hypertension secondary to other renal disorders, Hypertension secondary to endocrine disorders, Other secondary hypertension, Seconday hypertension, unspecified |
| Chronic Kidney Disease | 20002 | Non-cancer illness code, self-reported | 1192, 1193, 1519, 1520, 1607 | diabetic nephropathy, iga nephropathy, kidney nephropathy, renal failure requiring dialysis, renal failure |
| Chronic Kidney Disease | 41203  41205 | Diagnosis - main ICD9  Diagnoses - secondary ICD9 | 585,5859 | Chronic renal failure, Chronic renal failure |
| Chronic Kidney Disease | 41202  41204  40001  40002 | Diagnoses - main ICD10  Diagnoses – secondary ICD10  Underlying (primary) cause of death: ICD10  Contributory (secondary) cause of death: ICD10 | I12.0,I13.1,I13.2,N18,N18.0,N18.1,N18.2,N18.3,N18.4,N18.5,N18.8,N18.9 | Hypertensive renal disease with renal failure, Hypertensive heart and renal disease with renal failure, Hypertensive heart and renal disease with both (congestive) heart failure and renal failure, Chronic renal failure, End-stage renal disease, Chronic kidney disease, stage 1, Chronic kidney disease, stage 2, Chronic kidney disease, stage 3, Chronic kidney disease, stage 4, Chronic kidney disease, stage 5, Other chronic renal failure, Chronic renal failure, unspecified |
| Chronic Kidney Disease | 41200  41210 | Operative procedures – main OPCS  Operative procedures – secondary OPCS | M01,M01.1,M01.2,M01.3,M01.4,M01.5,M01.8,M01.9 | Transplantation of kidney, Autotransplantation of kidney, Allotransplantation of kidney from live donor, Allotransplantation of kidney from cadaver NEC, Allotransplantation of kidney from cadaver heart beating, Allotransplantation of kidney from cadaver heart non-beating, Other specified transplantation of kidney, Unspecified transplantation of kidney |
| Coronary artery disease | 20002 | Non-cancer illness code, self-reported | 1075 | heart attack/myocardial infarction |
| Coronary artery disease | 20004 | Operation code, self-reported | 1070,1095,1523 | coronary angioplasty (ptca) +/- stent, coronary artery bypass grafts (cabg), triple heart bypass |
| Coronary artery disease | 41203  41205 | Diagnosis - main ICD9  Diagnoses - secondary ICD9 | 410,4109,411,4119,412,4129 | Acute myocardial infarction, Other acute and subacute forms of ischaemic heart disease, Old myocardial infarction, Acute myocardial infarction, Other acute and subacute forms of ischaemic heart disease, Old myocardial infarction |
| Coronary artery disease | 41202  41204  40001  40002 | Diagnoses - main ICD10  Diagnoses – secondary ICD10  Underlying (primary) cause of death: ICD10  Contributory (secondary) cause of death: ICD10 | I21,I21.0,I21.1,I21.2,I21.3,I21.4,I21.9,I22,I22.0,I22.1, I22.8,I22.9,I23,I23.0,I23.1,I23.2,I23.3,I23.4,I23.5,I23.6, I23.8,I24,I24.0,I24.1,I24.8,I24.9,I25.2 | Acute myocardial infarction Acute transmural myocardial infarction of anterior wall, Acute transmural myocardial infarction of inferior wall, Acute transmural myocardial infarction of other sites, Acute transmural myocardial infarction of unspecified site, Acute subendocardial myocardial infarction, Acute myocardial infarction, unspecified, Subsequent myocardial infarction, Subsequent myocardial infarction of anterior wall, Subsequent myocardial infarction of inferior wall, Subsequent myocardial infarction of other sites, Subsequent myocardial infarction of unspecified site, Certain current complications following acute myocardial infarction, Haemopericardium as current complication following acute myocardial infarction, Atrial septal defect as current complication following acute myocardial infarction, Ventricular septal defect as current complication following acute myocardial infarction, Rupture of cardiac wall without haemopericardium as current complication following acute myocardial infarction, Rupture of chordae tendineae as current complication following acute myocardial infarction, Rupture of papillary muscle as current complication following acute myocardial infarction , Thrombosis of atrium, auricular appendage and ventricle as current complications following acute myocardial infarction, Other current complications following acute myocardial infarction, Other acute ischaemic heart diseases, Coronary thrombosis not resulting in myocardial infarction, Dressler's syndrome, Other forms of acute ischaemic heart disease, Acute ischaemic heart disease, unspecified, Old myocardial infarction |
| Coronary artery disease | 41200  41210 | Operative procedures – main OPCS  Operative procedures – secondary OPCS | K40,K40.1,K40.2,K40.3,K40.4,K40.8,K40.9,K41,K41.1,K41.2,K41.3,K41.4,K41.8,K41.9,K42,K42.1,K42.2,K42.3,K42.4,K42.8, K42.9,K43,K43.1,K43.2,K43.3,K43.4,K43.8,K43.9,K44,K44.1, K44.2,K44.8,K44.9,K45.1,K45.2,K45.3,K45.4,K45.5,K45.6,K45.8, K45.9,K46,K46.1,K46.2,K46.3,K46.4,K46.5,K49.2,K49.3,K49.4,K49.8,K49.9,K50.1,K50.2,K50.4,K75.1,K75.2, K75.3,K75.4,K75.8,K75.9 | Saphenous vein graft replacement of coronary artery, Saphenous vein graft replacement of one coronary artery, Saphenous vein graft replacement of two coronary arteries, Saphenous vein graft replacement of three coronary arteries , Saphenous vein graft replacement of four or more coronary arteries, Other specified saphenous vein graft replacement of coronary artery, Unspecified saphenous vein graft replacement of coronary artery, Other autograft replacement of coronary artery, Autograft replacement of one coronary artery NEC, Autograft replacement of two coronary arteries NEC, Autograft replacement of three coronary arteries NEC, Autograft replacement of four or more coronary arteries NEC, Other specified other autograft replacement of coronary artery, Unspecified other autograft replacement of coronary artery, Allograft replacement of coronary artery, Allograft replacement of one coronary artery, Allograft replacement of two coronary arteries, Allograft replacement of three coronary arteries, Allograft replacement of four or more coronary arteries, Other specified allograft replacement of coronary artery, Unspecified allograft replacement of coronary artery, Prosthetic replacement of coronary artery, Prosthetic replacement of one coronary artery, Prosthetic replacement of two coronary arteries, Prosthetic replacement of three coronary arteries, Prosthetic replacement of four or more coronary arteries, Other specified prosthetic replacement of coronary artery, Unspecified prosthetic replacement of coronary artery, Other replacement of coronary artery, Replacement of coronary arteries using multiple methods, Revision of replacement of coronary artery, Other specified other replacement of coronary artery, Unspecified other replacement of coronary artery, Double anastomosis of mammary arteries to coronary arteries, Double anastomosis of thoracic arteries to coronary arteries NEC, Anastomosis of mammary artery to left anterior descending coronary artery, Anastomosis of mammary artery to coronary artery NEC, Anastomosis of thoracic artery to coronary artery NEC, Revision of connection of thoracic artery to coronary artery, Other specified connection of thoracic artery to coronary artery, Unspecified connection of thoracic artery to coronary artery, Other bypass of coronary artery, Double implantation of mammary arteries into heart, Double implantation of thoracic arteries into heart NEC, Implantation of mammary artery into heart NEC, Implantation of thoracic artery into heart NEC, Revision of implantation of thoracic artery into heart, Other specified other bypass of coronary artery, Unspecified other bypass of coronary artery, Percutaneous transluminal balloon angioplasty of one coronary artery, Percutaneous transluminal balloon angioplasty of multiple coronary arteries, Percutaneous transluminal balloon angioplasty of bypass graft of coronary artery, Percutaneous transluminal cutting balloon angioplasty of coronary artery, Other specified transluminal balloon angioplasty of coronary artery, Unspecified transluminal balloon angioplasty of coronary artery, Percutaneous transluminal laser coronary angioplasty, Percutaneous transluminal coronary thrombolysis using streptokinase, Percutaneous transluminal atherectomy of coronary artery, Percutaneous transluminal balloon angioplasty and insertion of 1-2 drug-eluting stents into coronary artery, Percutaneous transluminal ba, Percutaneous transluminal balloon angioplasty and insertion of 1-2 stents into coronary artery, Percutaneous transluminal balloon angioplasty and insertion of 3 or more stents into coronary artery NEC, Other specified percutaneous transluminal balloon angioplasty and insertion of stent into coronary artery, Unspecified percutaneous transluminal balloon angioplasty and insertion of stent into coronary artery |
| Coronary artery disease | 20002 | Non-cancer illness code, self-reported | 1075 | heart attack/myocardial infarction |
| Myocardial | 42001 | Source of first myocardial infarction report | 0, 1, 2 | Self-reported only, hospital admission, death only (UK Biobank algorithm-defined phenotype) |
| Tobacco Use - Current | 20116 | Smoking status | 2 | Current |
| Tobacco Use - Previous | 20116 | Smoking status | 1 | Previous |
| Tobacco Use - Never | 20116 | Smoking status | 0 | Never |

#### **Table 2 Predictive performance of models for 5-year atrial fibrillation risk by subgroups of age**

| **AUROC** | | | | |
| --- | --- | --- | --- | --- |
|  | Age < 60 | Age 60-70 | Age > 70 | Overall |
| N | 13887 | 20725 | 14681 | 49293 |
| CHARGE-AF | 0.739 (0.678-0.798) | 0.666 (0.639-0.697) | 0.688 (0.657-0.719) | 0.785 (0.769-0.801) |
| AF PRS | 0.607 (0.524-0.678) | 0.617 (0.584-0.648) | 0.641 (0.612-0.672) | 0.618 (0.598-0.639) |
| ECG-AI | 0.634 (0.565-0.701) | 0.645 (0.616-0.679) | 0.687 (0.658-0.719) | 0.705 (0.686-0.724) |
| CHARGE + PRS | 0.752 (0.700-0.804) | 0.693 (0.659-0.721) | 0.734 (0.709-0.764) | 0.802 (0.786-0.818) |
| ECG-AI + PRS | 0.658 (0.582-0.728) | 0.674 (0.640-0.706) | 0.726 (0.700-0.754) | 0.728 (0.709-0.747) |
| CHARGE + ECG-AI | 0.738 (0.684-0.795) | 0.697 (0.665-0.726) | 0.732 (0.705-0.761) | 0.802 (0.786-0.818) |
| CHARGE + ECG-AI + PRS | 0.757 (0.692-0.813) | 0.715 (0.684-0.741) | 0.769 (0.738-0.793) | 0.817 (0.802-0.832) |
| **Average Precision** | | | | |
|  | Age < 60 | Age 60-70 | Age > 70 | Overall |
| CHARGE-AF | 0.016 (0.011-0.030) | 0.050 (0.040-0.067) | 0.073 (0.063-0.088) | 0.053 (0.048-0.061) |
| AF PRS | 0.010 (0.0068-0.016) | 0.037 (0.030-0.050) | 0.086 (0.072-0.11) | 0.038 (0.028-0.045) |
| ECG-AI | 0.054 (0.020-0.13) | 0.063 (0.046-0.092) | 0.15 (0.12-0.20) | 0.085 (0.071-0.11) |
| CHARGE + PRS | 0.019 (0.012-0.033) | 0.074 (0.055-0.11) | 0.095 (0.079-0.13) | 0.053 (0.048-0.061) |
| ECG-AI + PRS | 0.052 (0.021-0.13) | 0.079 (0.052-0.12) | 0.16 (0.13-0.22) | 0.095 (0.079-0.12) |
| CHARGE + ECG-AI | 0.076 (0.030-0.16) | 0.081 (0.059-0.12) | 0.15 (0.11-0.24) | 0.098 (0.081-0.13) |
| CHARGE + ECG-AI + PRS | 0.067 (0.026-0.15) | 0.099 (0.065-0.14) | 0.18 (0.12-0.27) | 0.11 (0.091-0.15) |
| Event rate | 0.00634 | 0.0221 | 0.05 | 0.0239 |

#### **Table 3 Predictive performance of models for 5-year atrial fibrillation risk by subgroups of sex**

| **AUROC** | | | |
| --- | --- | --- | --- |
|  | Men | Women | Overall |
| N | 23551 | 25742 | 49293 |
| CHARGE-AF | 0.757 (0.737-0.779) | 0.788 (0.761-0.815) | 0.785 (0.769-0.801) |
| AF PRS | 0.620 (0.592-0.644) | 0.619 (0.580-0.656) | 0.618 (0.598-0.639) |
| ECG-AI | 0.707 (0.682-0.733) | 0.666 (0.634-0.699) | 0.705 (0.686-0.724) |
| CHARGE + PRS | 0.779 (0.759-0.802) | 0.802 (0.772-0.831) | 0.802 (0.786-0.818) |
| ECG-AI + PRS | 0.729 (0.706-0.753) | 0.695 (0.662-0.728) | 0.728 (0.709-0.747) |
| CHARGE + ECG-AI | 0.788 (0.768-0.808) | 0.791 (0.763-0.823) | 0.802 (0.786-0.818) |
| CHARGE + ECG-AI + PRS | 0.805 (0.786-0.823) | 0.806 (0.779-0.834) | 0.817 (0.802-0.832) |
| **Average Precision** | | | |
|  | Men | Women | Overall |
| CHARGE-AF | 0.062 (0.054-0.073) | 0.035 (0.029-0.048) | 0.053 (0.048-0.061) |
| AF PRS | 0.055 (0.046-0.066) | 0.025 (0.021-0.033) | 0.038 (0.028-0.045) |
| ECG-AI | 0.11 (0.088-0.15) | 0.042 (0.030-0.068) | 0.085 (0.071-0.11) |
| CHARGE + PRS | 0.079 (0.067-0.097) | 0.043 (0.035-0.058) | 0.053 (0.048-0.061) |
| ECG-AI + PRS | 0.13 (0.099-0.17) | 0.048 (0.034-0.075) | 0.095 (0.079-0.12) |
| CHARGE + ECG-AI | 0.12 (0.095-0.16) | 0.055 (0.042-0.079) | 0.098 (0.081-0.13) |
| CHARGE + ECG-AI + PRS | 0.14 (0.11-0.19) | 0.060 (0.049-0.834) | 0.11 (0.091-0.15) |
| Event rate | 0.0332 | 0.0154 | 0.0239 |

#### **Table 4 Increase in discrimination for 5-year risk of incident atrial fibrillation**

| **Model** | **Improvement in AUROC (95% CI)** | **Improvement in AP (95% CI)** |
| --- | --- | --- |
| Adding PRS to CHARGE | 0.017 (0.0096-0.025) | 0.0136 (0.00846-0.0210) |
| Adding PRS to ECG-AI | 0.023 (0.012-0.034) | 0.0102 (0.00461-0.0186) |
| Adding ECG-AI to CHARGE | 0.017 (0.0091-0.026) | 0.0449 (0.0299-0.0732) |
| Adding PRS to ECG-AI and CHARGE | 0.015 (0.0089-0.021) | 0.0139 (0.00493-0.0217) |

#### **Table 5 Net reclassification of incident AF risk using Predict-AF3 versus two-component models**

|  |  | **Predict-AF3** | |  | **Predict-AF3** | |
| --- | --- | --- | --- | --- | --- | --- |
|  | **CHARGE ECG** | ≤2.5% | >2.5% | **CHARGE PRS** | ≤2.5% | >2.5% |
| **Cases** | ≤2.5% | 198 | 61 | ≤2.5% | 193 | 51 |
|  | >2.5% | 37 | 529 | >2.5% | 42 | 539 |
| **Non-Cases** | ≤2.5% | 30172 | 2134 | ≤2.5% | 29829 | 1764 |
|  | >2.5% | 2775 | 13387 | >2.5% | 3118 | 13757 |
|  | NRI | 0.039 (0.015-0.066) | | NRI | 0.033 (0.0082-0.059) | |
|  | NRI+ | 2.59% (0.18-5.3) | | NRI+ | 0.52% (-2.06-3.00) | |
|  | NRI- | 1.35% (1.06-1.67) | | NRI- | 2.81% (2.54-3.08) | |
|  | Green cells denote appropriate reclassification  Red cells denote inappropriate reclassification  NRI=Net reclassification improvement; NRI+=Event reclassification improvement; NRI-=Non-event reclassification improvement  NRI values calculated using the Kaplan-Meier estimator to account for censored survival data^14^ | | | | | |

### Supplementary Figures

#### **Figure 1: Calibration of incident atrial fibrillation**

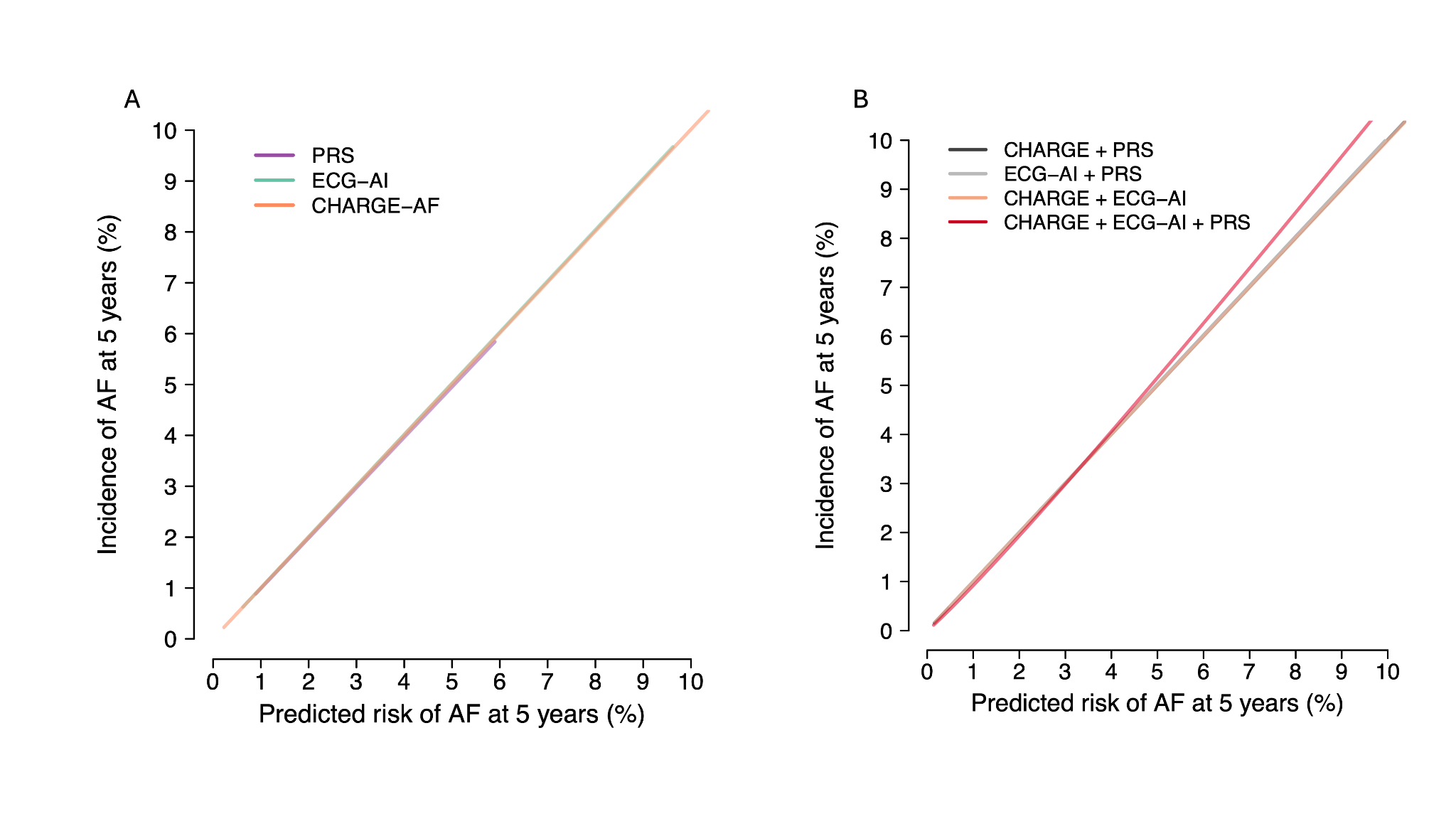

In Panel A, model calibration using a polygenic risk score (PRS; teal) versus an ECG-derived artificial intelligence (ECG-AI) prediction (turquoise) and the CHARGE-AF score depicted is, where the x-axis depicts predicted atrial fibrillation (AF) risk at 5 years, and the y-axis depicts observed heart failure incidence. Curves were fit using adaptive hazard regression. Panel B depicts model calibration using models a) combining polygenic risk score (PRS) and CHARGE-AF (black), a model combining an ECG-derived artificial intelligence (ECG-AI) prediction and the PRS (grey), a model combining ECG-AI and the CHARGE-AF (orange), and a model combining ECG-AI, CHARGE-AF and the PRS (Predict-AF3, red)
